# A biomarker catalog for inflammatory bowel disease medications

**DOI:** 10.1101/2025.10.22.25338516

**Authors:** T. J. Hart, Kiana A. West, António José Preto, Daniel Domingo-Fernández

**Author notes:** Corresponding author. Daniel Domingo Fernández. Enveda Therapeutics, Inc., Boulder, CO, USA. These authors contributed equally to this work (António José Preto, Daniel Domingo-Fernández).

## Abstract

**Introduction:** Inflammatory bowel disease (IBD), which includes Crohn’s disease (CD) and ulcerative colitis (UC), is a multifaceted illness with diverse manifestations. Current treatment strategies are inadequate, with low response rates, secondary loss of response, and serious side effects.

**Methods:** We leveraged a large multiomics dataset (SPARC IBD) to compile a catalog of biomarkers for (i) disease severity, (ii) medication use, and (iii) response to a medication. Data were stratified based on IBD subtype (UC and CD) and tissue type; biomarker associations were identified using mixed-effects models to account for multiple medication groups, disease status (e.g., remission, active, etc.), and repeated sampling.

**Results:** Our catalog lists thousands of potential biomarkers for disease severity (9,657), medication effect (293), and medication response (59). These biomarkers, both known and novel, span multiple diagnoses, tissues, and medications.

**Conclusion:** These therapy-specific signatures will enable tracking of treatment response, diagnosis of disease severity, and identification of new therapeutic targets.

## 1. Introduction

Inflammatory bowel disease (IBD), which includes Crohn’s disease (CD) and ulcerative colitis (UC), presents with complex clinical heterogeneity. Patients exhibit variation in symptoms, affected anatomy, disease behavior, and therapeutic outcomes (Imhann *et al*., 2019). Although numerous medications exist, roughly one-third of patients fail to reach clinical remission with initial therapy, and nearly half experience secondary loss of response over time (Gibble *et al*., 2023; Kumar *et al*., 2024). This pattern frequently leads to a series of therapy changes in a trial□and□error manner, exposing patients to prolonged inflammation and heightened risks of complications such as strictures, fistulas, or the need for surgery (Berg *et al*., 2019). Although the molecular basis of this therapeutic variability remains poorly understood, it is believed to stem from distinct immunological and molecular endotypes that underlie treatment resistance or response within specific patient subgroups (Selin *et al*., 2021). Consequently, there is an urgent need to align treatment choice with underlying biological drivers, focusing on patient population-specific profiles and broadening precision medicine use in IBD (Noor *et al*., 2022).

Researchers are increasingly leveraging transcriptomic and proteomic profiling to address the heterogeneity of IBD and improve the prediction of treatment outcomes. Unlike static genomic markers, these high-throughput ‘omics’ capture dynamic snapshots of the inflammatory state. For instance, the tissue transcriptome reveals active immune pathways while the circulating proteome reflects downstream effector molecules such as cytokines. Both modalities can thus directly monitor disease activity and therapeutic effects, providing sensitive biomarkers that change throughout the patient journey. Recent multi-omic studies underscore this potential. In the large prospective SPARC IBD cohort, our previous work demonstrated that leveraging multiple omics enabled the accurate classification of CD and UC patients (Preto *et al*., 2025). Furthermore, we identified patient subgroups with distinct inflammation profiles.

Similarly, a prospective multi-omics study that combined stool metagenomics with serum metabolomics and proteomics improved accuracy in predicting biologic therapy responses, far exceeding clinical predictors alone (Lee *et al*., 2021). However, while multiple recent multi-omics studies have begun to explore the prediction of treatment outcomes (Mishra *et al*., 2022; Mehta *et al., 2023*; Schirmer *et al*., 2024; Hassan-Zahraee *et al*., 2024; Chen *et al*., 2025), comprehensive and clinically validated biomarkers remain lacking, leaving a critical knowledge gap in understanding and predicting heterogeneity in drug response.

Despite increasing use of advanced therapies for IBD, no clinically validated molecular biomarkers can predict which patients will respond to a specific treatment (Atreya and Neurath, 2024). Drug selection remains empirical, mainly guided by disease phenotype, prior exposure, and physician preference rather than individualized biology (Juillerat *et al*., 2022). This lack of precision contributes to suboptimal outcomes and delayed disease control, particularly in patients who must cycle through multiple therapies before achieving remission.

The present study aims to identify medication group signatures in CD and UC patients using transcriptomic and proteomic data from the SPARC IBD cohort. We first identified signatures that differentiate between patients in remission and those with active disease while on the same medication. Next, we focused on unique signatures in remission patients for six medication families (e.g., TNF-α inhibitors, JAK inhibitors), revealing genes and proteins regulated by each medication. Lastly, we investigated signatures that were associated with active disease and reversed by a particular medication. The biomarker catalog will enable prompt confirmation of treatment effectiveness, reducing the prescription of ineffective therapies. By integrating therapy-matching and severity-grading biomarkers, this study seeks to accelerate precision-trial design and personalize IBD care.

## 2. Methods

### 2.1. SPARC IBD cohort

In this study, we leverage data from the SPARC IBD cohort, downloaded as of May 2025 (https://www.crohnscolitisfoundation.org/research/plexus/sparc-ibd) (Raffals *et al*., 2022). We focused on patients diagnosed with either CD or UC **(Figure 1A)**, excluding samples without a definitive diagnosis. We extracted relevant metadata **(Supplementary Table 1)** and medication information for these individuals (**see Section 2.2**) as well as disease severity (mild, moderate, severe, and remission). The distribution of the most relevant metadata for the samples used in the study is displayed in **Supplementary Figure 1**. Multiple sample types were collected from each patient at different time points. We utilized transcriptomics from gut biopsies (colon or small intestine) and proteomics from plasma.

**Table 1.**
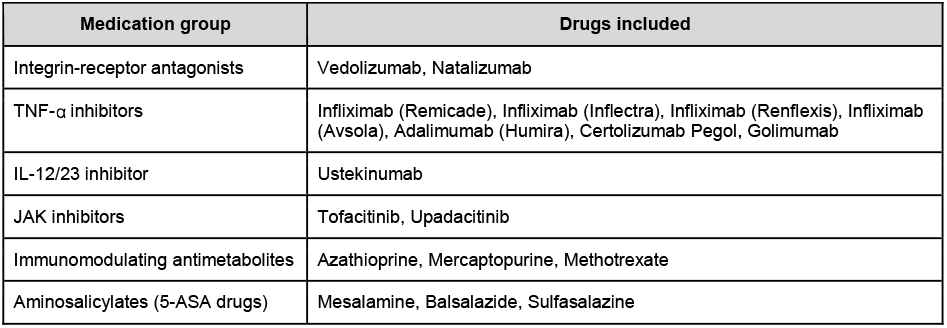
Medication families and their corresponding drugs.

**Figure 1.**
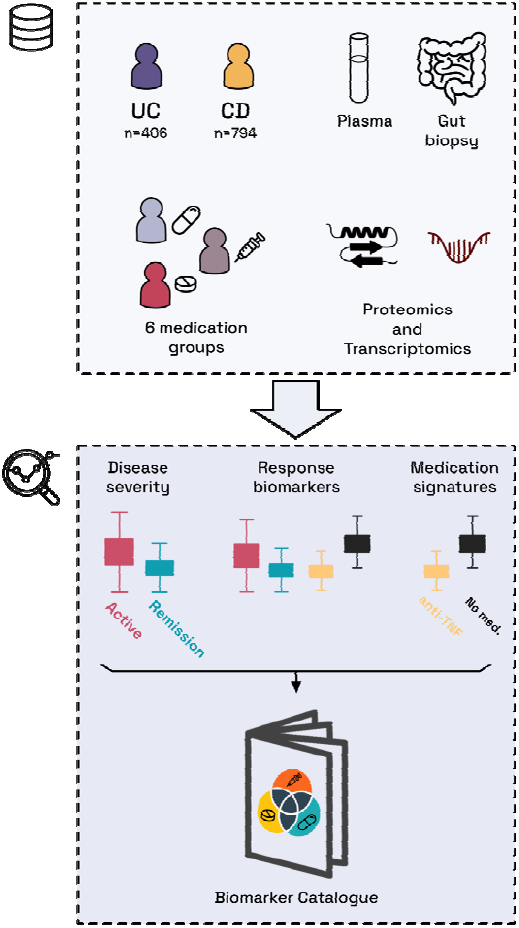
Graphical representation of this work. **Upper box)** We leverage proteomics and transcriptomics from plasma and gut biopsies from UC and CD patients treated with six different medications. **Bottom box)** We investigate signatures specific to each medication, disease severity, and medication responder biomarkers.

### 2.2. Grouping individual therapies into medication families

The landscape of IBD drugs is vast. In our dataset, we have a total of 18 unique drugs which we grouped into medication groups that share an MOA or target(s), such as TNF-α inhibitors **(Table 1). Supplementary Figure 2** shows the number of patients for each medication group. In addition, we include patients who have no medication to use as a baseline in our analyses and modeling.

### 2.3. Data processing

Overall, the transcriptomics data preprocessing workflow is similar to what we used in our previous publication for this dataset (Preto et al., 2025), and more details are available in **Supplementary Text 2**.

The proteomics data is Olink®, and it is derived from plasma samples. Since proteomics data is already processed and does not have a strong batch effect, we only filtered out samples with a quality control warning tag. The total number of proteomics features remaining is 2,939. These features encompass multiple measurements of the same protein across different panels. Lastly, we log2-transformed transcriptomics before modelling; we note that Olink NPX values are already log2-transformed.

### 2.4. Data modeling

In light of the samples’ biological separation, we split the dataset into strata based on UC and CD, and, within each of these, by tissue type (transcriptomics only). We utilized disease severity, defined by Mayo scores, and categorized it into i) active (including moderate and severe patients), and ii) remission. As mild samples are not different enough from moderate or remission, we ignored them in these analyses. To isolate the effects of a given drug from the effects of other medications, we filtered out any patient who switched from one drug to another or was on multiple medications.

For each of the following analyses, we define a comparison of two groups (within a stratum) of samples on a transcript/protein as statistically significant if it has an adjusted *p*-value (Benjamini-Hochberg corrected) below 0.05. For the comparison to be considered as significantly up- or down-regulated, it must have an absolute log2 fold change exceeding 1.0 for transcriptomics (a 100% increase/decrease) and 0.5 for proteomics (∼40% increase/decrease).

#### 2.4.1. Identifying medication-specific signatures correlated with disease severity

First, we split the data into two groups: remission samples and active samples, where active samples comprise samples with moderate and severe disease severity. We filtered these two subgroups by individual medication families to minimize variability from different medication effects.

We conducted this comparison using mixed-effects models, which allow us to consider the patient identifiers as a random effect and the disease severity as a fixed effect, where remission samples are the reference. The model output indicates the change (coefficient) for a protein/transcript associated with active disease. These coefficients represent fold changes between different comparisons, as our variables of interest are categorical.

##### Identifying medication-specific signatures in remission patients

To identify medication-specific biomarkers in remission samples, we compared patients on individual medication families with non-medicated patients. We use mixed-effects models to compute the comparisons. Similar to 2.4.1, we considered the patients as random effects. This time, using remission patients only, we considered the medication group as a fixed effect and used the non-medicated samples as the reference. The model output here indicates the changes (fold change coefficients) for medicated patients against the reference as the effect (e.g., medication group X vs. non-medication, medication group Y vs. non-medication, etc.).

##### Identifying medication response biomarkers

Since the IBD Plexus cohort lacks sufficient longitudinal follow-up (e.g., paired pre-/post-treatment samples), the traditional concept of within-patient medication effect is not viable. Instead, the large sample size enables a cross-sectional strategy by comparing sample groups.

We define medication response biomarkers **(Figure 2)** based on two interrelated criteria:

1. Significantly up- or down-regulated in active compared to remission patients on the given medication (Section 2.4.1).
2. If (1) is fulfilled, the biomarker must be significantly and oppositely regulated in remission samples on a given medication compared to those not on medication (Section 2.4.2).

**Figure 2.**
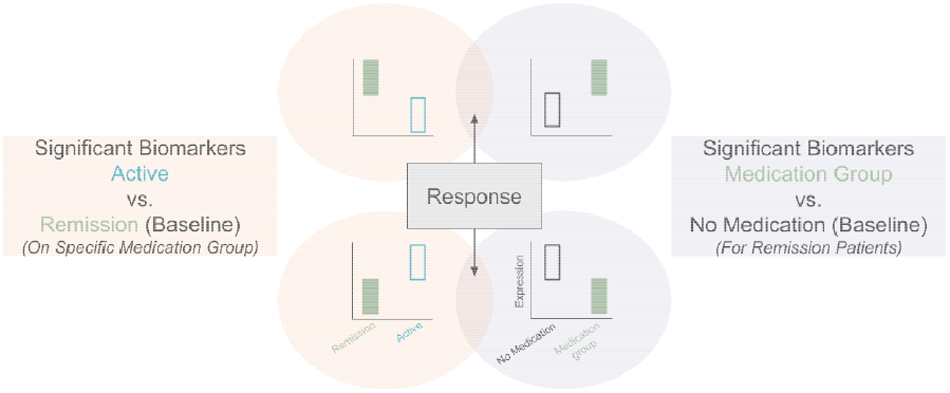
Definitions for medication response biomarkers.

From a clinical perspective, these biomarkers could facilitate confirmation of treatment effectiveness for patients.

### 2.5. Implementation

We implemented the scripts and notebook in Python version 3.10 and managed them using Poetry (https://python-poetry.org/). We leveraged open-source libraries such as NumPy (Harris *et al*., 2020), Pandas (McKinney, 2010), and SciPy (Virtanen et al., 2020). We employed Seaborn (Waskom, 2021) and Matplotlib (Hunter, 2007) for visualization. The mixed-effects models are implemented using statsmodels (Seabold and Perktold, 2010).

## 3. Results

### 3.1. Medication signatures based on disease severity

We begin our analysis by identifying biomarkers (proteins and transcripts) that are different between active and remission (baseline) patients receiving the same medication group (e.g., TNF-α inhibitors) within each sample subset (e.g., CD-small intestine). To do this, we compared samples from remission patients in a medication group with samples from patients in the same medication group but with active disease.

In the proteomics results **(Figure 3A)**, we notice that UC has the strongest signal, and 5-ASA drug patients have the most significant number of regulated protein measurements (1,035) for the UC group, followed by TNF-α inhibitors (578). In CD, the immunomodulators are the only medication group with significant changes (in 17 protein measurements).

**Figure 3.**
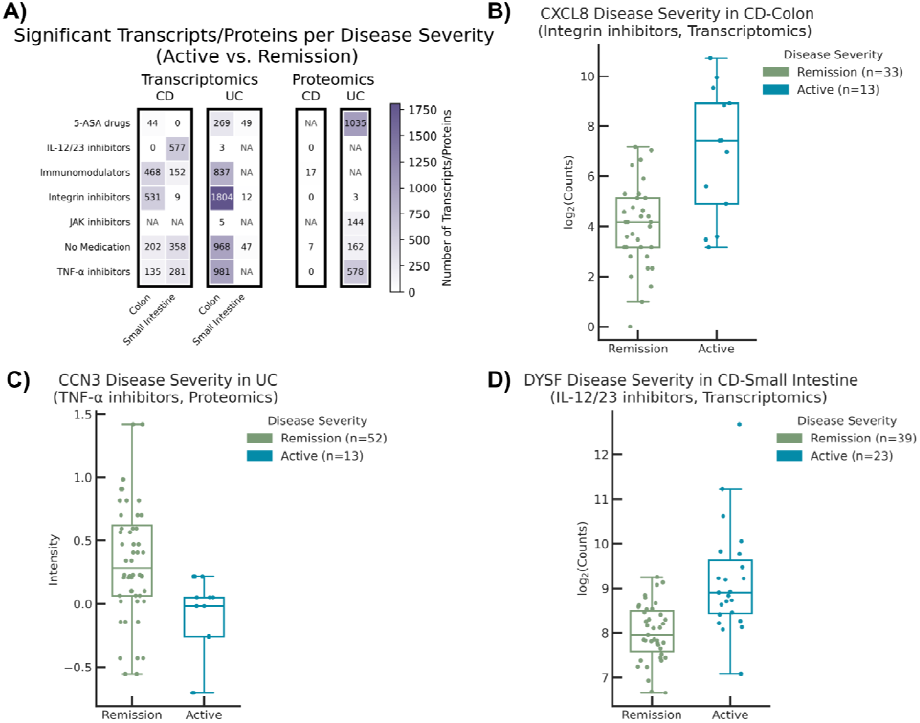
**A)** Number of significantly (adjust-*p*-value < 0.05) regulated transcripts/proteins in the active vs. remissio (baseline) comparison for each medication group across all subpopulations. NA indicates insufficient samples t conduct the analyses, and zero indicates no significant genes for that particular group. The counts for the proteomics are the number of significant protein measurements. **B-D** Examples of significantly regulated genes/proteins for different modalities (**Transcriptomics threshold:** absolute fold change > **1**; **Proteomics threshold:** absolute fol change > 0.5).

In the transcriptomics results, we notice that UC has the strongest signal; however, it is confined to the colon because there is little signal in UC-small intestine, as expected. Notably, in UC-colon, integrin inhibitors have the highest number of regulated transcripts (1,804), followed by TNF-α inhibitors (981). In CD, both tissues provide an adequate signal, reflecting the distribution of disease activity across the digestive tract. Colon is most affected by integrin inhibitors (531) and Immunomodulators (468); the small intestine is most affected by the following medications: IL-12/23 inhibitors (577) and TNF-α inhibitors (281).

**Figure 3 (B-D)** shows examples of regulated markers for each omics type. All three examples clearly separate the two groups (active and remission). We performed pathway analysis on these sets of regulated transcripts and saw that, for patients in CD-colon and small intestine subgroups on TNF-α inhibitors, many of the affected pathways were inflammation- or immune-related (see rem_vs_act_transcriptomics.xlsx in **Supplementary Data**).

### 3.2. Medication signatures in remission patients

Here, we aim to understand the changes each medication group induces in patients in remission for both UC and CD. To do this, we compare samples of remission patients taking a specific medication group (e.g., TNF-α inhibitors) with those who do not receive any medication but are also in remission **(Figure 4A)**. This exercise allows us to identify which proteins and transcripts are regulated by each medication without the disease status’s impact.

**Figure 4.**
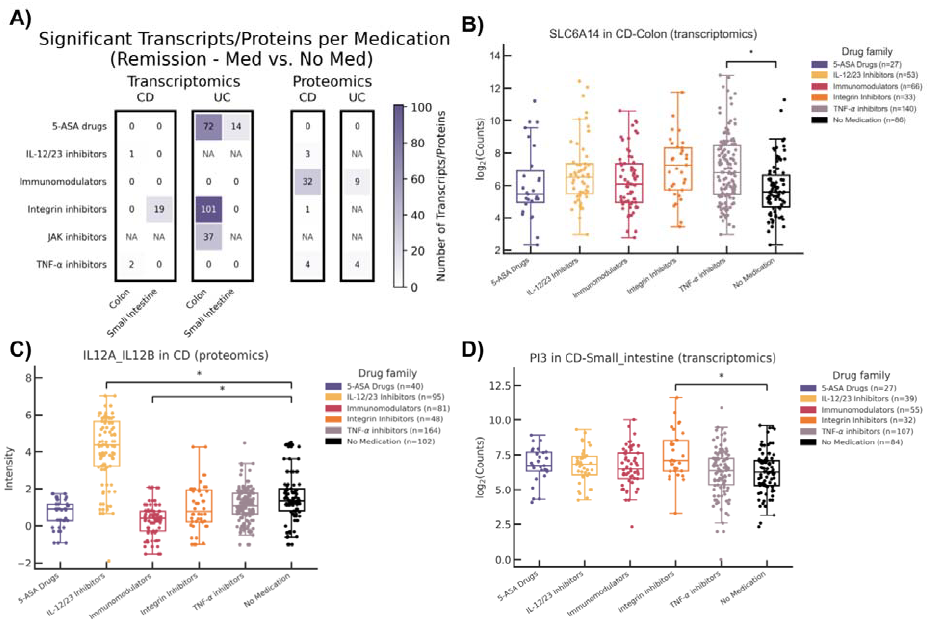
**A)** Number of significantly (adjust-*p*-value < 0.05) regulated transcripts/proteins for the medication vs no medication comparison for each medication group across all subgroups. NA indicates insufficient samples to conduct the analyses, and zero indicates no significant genes for that particular group. The counts for the proteomics are th number of significant protein measurements. **B-D** Examples of significant genes/proteins for different modalities. * indicates the significant groups with respect to no medication (**Transcriptomics threshold:** absolute fold change > **1**; **Proteomics threshold:** absolute fold change > 0.5). The gene names of all these significantly regulated proteins/transcripts are available in the **Supplementary Data** (workbook name: catalog_workbook_2_med_vs_no_med.xlsx).

In UC proteomics (**Figure 4B**), we observe that Immunomodulators and TNF-α inhibitors have the greatest number of significant marker measurements. In addition, it is important to note that Integrin inhibitors and IL-12/23 inhibitors did not have enough patients for this given subpopulation to be included in the analysis. Within the significant immunomodulator proteins, some noteworthy up-regulated genes are IL15 (Liu *et al*., 2000) and CXCL16 (Li *et al*., 2008), and some down-regulated proteins are FASLG (Souza *et al*., 2004) and CD160 (Piotrowska *et al*., 2021), all related to inflammation responses. Similarly, all four significant results in TNF-α inhibitors correspond to TNF measured in four different Olink panels, consistent with previous findings where TNF-α inhibitors have been shown to up-regulate TNF in the bloodstream (Berkhout *et al*., 2019). It is important to note that these same four protein measurements are also significant in the CD proteins, as seen in **Figure 4B** for TNF.

Compared to UC, CD proteomics includes more samples (**Figure 4B**), allowing us to model a few additional drugs. We identified significant protein measurements for IL-12/23 inhibitors (3), immunomodulators (32), integrin inhibitors (1), and TNF-α inhibitors (4). However, no significant proteins were found for 5-ASA drugs. Immunomodulators show a notably higher count of significant protein measurements for CD (32) compared to UC (9). This disparity might be attributed to the fact that while many immunomodulators are effective for both UC and CD, some, such as methotrexate, are only effective for CD (Raine *et al*., 2022).

Additionally, we investigated the transcriptomics signatures for the same medication families (**Figure 4A**). On a high level, transcriptomics and proteomics express differently depending on the diagnosis. For example, integrin inhibitors (101) and 5-ASA drugs (72) show the most statistically significant and regulated UC-Colon transcripts. Despite sufficient sample sizes, immunomodulators exhibit no significant transcriptomic findings across any subpopulations, directly contrasting their significant presence in all subpopulations of the proteomics data. For CD transcriptomics, the colon subgroup contained three significant transcripts: one for IL-12/23 inhibitors and two for TNF-α inhibitors. In the CD small intestine group, all 19 transcripts were associated with integrin inhibitors.

In summary, when searching for medication signatures, we see little signal in CD, except for integrin inhibitors in small intestine transcriptomics and immunomodulators in proteomics. Conversely, the UC-Colon transcripts are the most signal-rich group. Independent of diagnosis, the TNF-α inhibitors have low signal in both transcriptomics and proteomics. We catalog these medication-specific signatures in a complete list within **Supplementary Data**.

### 3.3. Response biomarkers per medication group

When overlapping the insights from disease severity and medication, we find the signal for regulated transcripts/proteins is much stronger when comparing active vs. remission medicated patients (**Figure 3A**) than medicated vs. non-medicated remission patients (**Figure 4A**). To deepen our understanding of how disease activity and medication interplay, we explored changes that the disease triggers and that the medication can reverse (section 3.3).

We observe just above 50 response biomarkers across all groups and modalities **(Figure 5A). Table 2** contains all the response biomarkers. 17.65% of response transcripts are from the Solute Carrier Family (SLC), suggesting that cellular transport mechanisms are possibly tied to medication response (Wojtal *et al*., 2009). Interestingly, seven of the nine medication response transcripts for 5-ASA drugs in UC colon show trends similar to those found with integrin inhibitors (indicated in bold in **Table 2**).

**Table 2.**
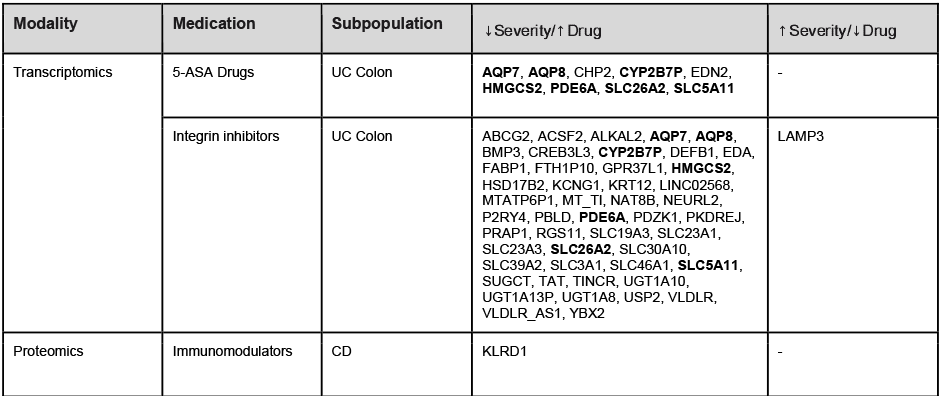
Response biomarker split by medication and group. Bold biomarkers are present in more than one group.

**Figure 5.**
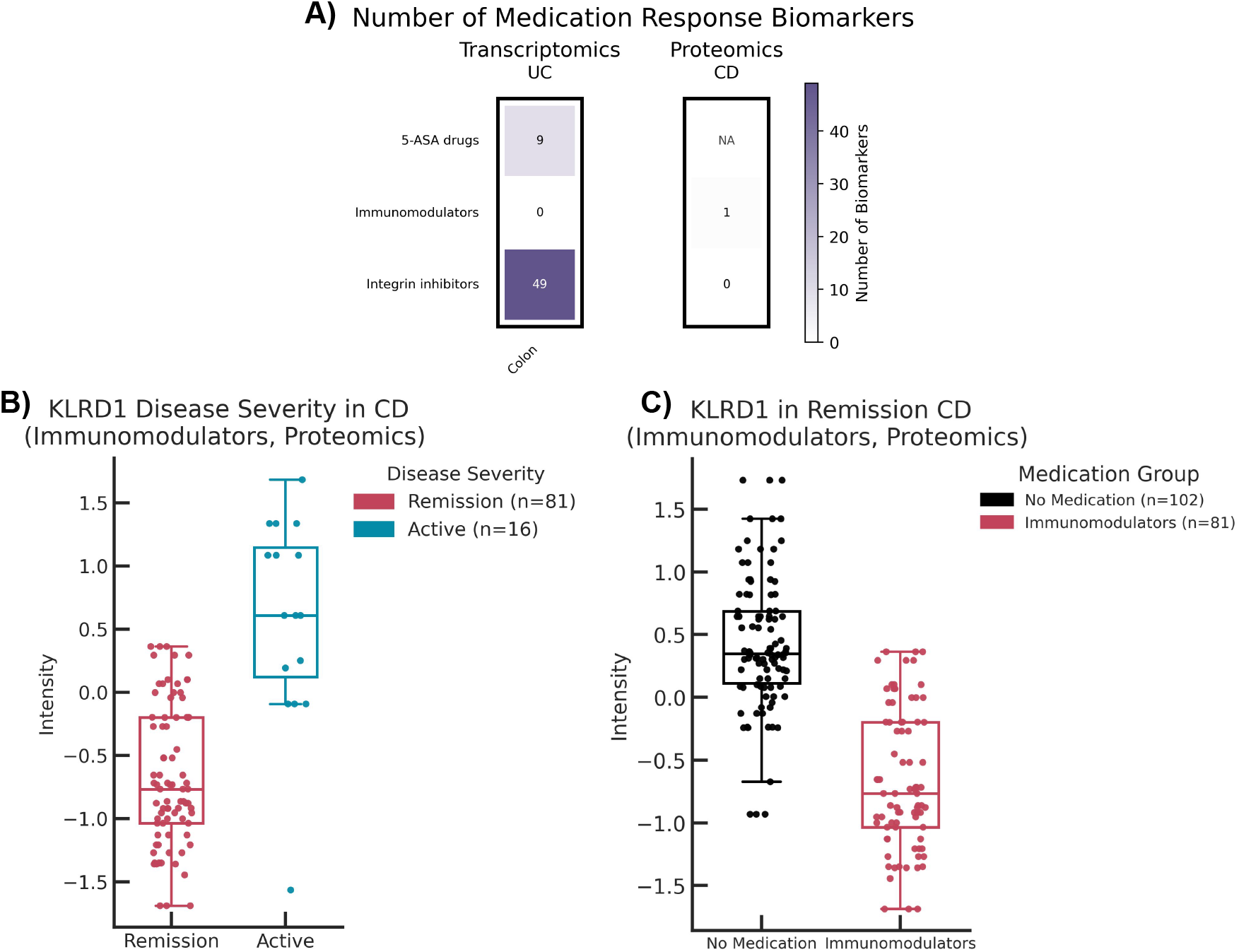
**A)** Number of medication response biomarkers for each medication group across all subgroups. NA indicates insufficient samples to conduct the analyses, and zero indicates no response biomarkers for that particular group. **B-C)** An example of an immunomodulator response protein, KLRD1. KLRD1 is a response protein because it is significantly down-regulated in active vs. remission (baseline) comparison, and is significantly up-regulated in the medication vs. no medication (baseline) comparison. Note the points for remission in **B** are the same points as those labeled as Immunomodulators in **C**.

All but one of the response transcripts are upregulated when comparing medication to no medication, and downregulated in active patients receiving a specific drug. This observation is likely because in our data, drugs tend to be associated with upregulation of transcripts **(Supplementary Figure 3)**. Finally, only one response biomarker, KLRD1 for immunomodulators in CD, was identified via proteomics **(Figure 5B and 5C)**.

It is important to recognize that specific drug-relevant proteins, initially identified as significant in medication vs. no-medication (section 3.2), are not response proteins. For example, TNF and IL12B are significantly up-regulated in medicated remission patients compared to those without medication (section 3.2), but they are not classified as response proteins. This is because they may not be disease-specific, i.e. their expression levels are not altered between remission and active states within their specific medication families. This is the case for a few other inflammation/immune-related genes as well: IL15, CD160, IL12A_IL12B, FCRL6, and NCR1. In the **Supplementary Data**, we also include proteins and transcripts that meet one of the criteria of response biomarker and not the other.

## 4. Discussion

Here, we have developed a biomarker catalog for IBD medications. Our study utilizes transcriptomic and proteomic data from the SPARC IBD cohort to pinpoint specific medication signatures, including new biomarkers for medication response. These proteo-transcriptomic signatures can serve a triple purpose: i) classify disease severity, ii) understand differential drug effects across disease subtypes and tissue types, and iii) identify medication responsiveness. Importantly, these biomarkers have the potential to enable non-invasive monitoring of disease activity across different medications, similar to established markers like C-reactive protein (CRP) and fecal calprotectin (FCP) (Langhorst *et al*., 2008). This catalog will allow for more rapid confirmation of treatment effectiveness and help guide patients away from ineffective therapies.

Despite using one of the most comprehensive datasets available, this work has a few limitations associated with IBD clinical data. Firstly, certain groups and medication families end up with insufficient samples to conduct any meaningful analysis. Secondly, unlike other studies focused on profiling changes before and after treatment (Lee *et al*., 2021), our dataset lacks in depth relative to what it has in breadth, since the number of data points per patient is too limited for longitudinal analysis. Thirdly, although our medication grouping (by MOA or target, e.g., TNF-α inhibitors) increased the study’s statistical power (by increasing sample size), it might mask subtle distinctions between drugs within the same therapeutic class. Finally, because our analyses were based on multiple subpopulations, we lacked sufficient patient numbers to include gender in the analyses, which could have further obscured distinctions.

Looking forward, the medication biomarkers identified here offer guidance for future work involving statistical or machine learning models involving IBD medication data. These therapy-specific signatures offer valuable potential for clinical application to (i) flag treatment response, (ii) diagnose disease severity, and (iii) propose potential new targets. This aligns with the broader aim of accelerating precision-trial design and personalizing routine IBD care. Furthermore, we foresee the inclusion of novel omics modalities like metabolomics to enrich our understanding of treatment response. While our current study did not investigate combination therapy signatures due to limited sample sizes, the SPARC IBD cohort’s continued growth may allow for this work’s extension to include such analyses.

## Acknowledgments

The results published here are in whole from the Study of a Prospective Adult Research Cohort with IBD (SPARC IBD). SPARC IBD is a component of the Crohn’s & Colitis Foundation’s IBD Plexus data exchange platform. SPARC IBD enrolls patients with an established or new diagnosis of IBD from sites throughout the United States and links data collected from the electronic health record and study-specific case report forms. Patients also provide blood, stool, and biopsy samples at selected times during follow-up. The design and implementation of the SPARC IBD cohort have been previously described.

We would like to thank David Healey for his feedback and comments on the manuscript. Magazine free icon by Nadiinko: Flaticon.com

## Author’s contributions

KAW, AJP, and DDF designed the study. AJP and TJH prepared the datasets. TJH analyzed the datasets. All authors interpreted the results and wrote the paper. All authors reviewed the manuscript. All authors have read and approved the final manuscript.

## Data and code availability

SPARC IBD is available upon approved application to Crohn’s & Colitis Foundation IBD Plexus (https://www.crohnscolitisfoundation.org/ibd-plexus). We released the scripts and notebooks to reproduce the work at https://github.com/enveda/ccf-medication.

## References

1. Atreya, R., and Neurath, M. F. (2024). Biomarkers for personalizing IBD therapy: the quest continues. Clinical Gastroenterology and Hepatology, 22(7), 1353–1364. 10.1016/j.cgh.2024.01.026

2. Berg, D. R., Colombel, J. F., and Ungaro, R. (2019). The role of early biologic therapy in inflammatory bowel disease. Inflammatory bowel diseases, 25(12), 1896–1905. 10.1093/ibd/izz059

3. Berkhout, L. C., l’Ami, M. J., Ruwaard, J., Hart, M. H., Heer, P. O. D., Bloem, K., et al. (2019). Dynamics of circulating TNF during adalimumab treatment using a drug-tolerant TNF assay. Science translational medicine, 11(477), eaat3356. 10.1126/scitranslmed.aat3356

4. Chen, L., Zhang, C., Niu, R., Xiong, S., He, J., Wang, Y., et al. (2025). Multi-Omics Biomarkers for Predicting Efficacy of Biologic and Small□Molecule Therapies in Adults With Inflammatory Bowel Disease: A Systematic Review. United European Gastroenterology Journal, 13(4), 517–530. 10.1002/ueg2.12720

5. Gibble, T. H., Naegeli, A. N., Grabner, M., Isenberg, K., Shan, M., et al. (2023). Identification of inadequate responders to advanced therapy among commercially-insured adult patients with Crohn’s disease and ulcerative colitis in the United States. BMC gastroenterology, 23(1), 63. 10.1186/s12876-023-02675-w

6. Harris, C. R., Millman, K. J., Van Der Walt, S. J., Gommers, R., Virtanen, P., Cournapeau, D., et al. (2020). Array programming with NumPy. Nature, 585(7825), 357–362. 10.1038/s41586-020-2649-2

7. Hassan-Zahraee, M., Ye, Z., Xi, L., Dushin, E., Lee, J., Romatowski, J., et al. (2024). Baseline serum and stool microbiome biomarkers predict clinical efficacy and tissue molecular response after ritlecitinib induction therapy in ulcerative colitis. Journal of Crohn’s and Colitis, 18(9), 1361–1370. 10.1093/ecco-jcc/jjad213

8. Hunter, J. D. (2007). Matplotlib: A 2D graphics environment. Computing in science & engineering, 9(03), 90–95.

9. Imhann, F., Van der Velde, K. J., Barbieri, R., Alberts, R., Voskuil, M. D., Vich Vila, A., and Weersma, R. K. (2019). The 1000IBD project: multi-omics data of 1000 inflammatory bowel disease patients; data release 1. BMC gastroenterology, 19(1), 5. 10.1186/s12876-018-0917-5

10. Juillerat, P., Grueber, M. M., Ruetsch, R., Santi, G., Vuillemoz, M., and Michetti, P. (2022). Positioning biologics in the treatment of IBD: A practical guide–Which mechanism of action for whom?. Current research in pharmacology and drug discovery, 3, 100104. 10.1016/j.crphar.2022.100104

11. Kumar, M., Murugesan, S., Ibrahim, N., Elawad, M., and Al Khodor, S. (2024). Predictive biomarkers for anti-TNF alpha therapy in IBD patients. Journal of Translational Medicine, 22(1), 284. 10.1186/s12967-024-05058-1

12. Langhorst, J., Elsenbruch, S., Koelzer, J., Rueffer, A., Michalsen, A., and Dobos, G. J. (2008). Noninvasive markers in the assessment of intestinal inflammation in inflammatory bowel diseases: performance of fecal lactoferrin, calprotectin, and PMN-elastase, CRP, and clinical indices. Official journal of the American College of Gastroenterology| ACG, 103(1), 162–169.

13. Lee, J. W. J., Plichta, D., Hogstrom, L., Borren, N. Z., Lau, H., Gregory, S. M., et al. (2021). Multi-omics reveal microbial determinants impacting responses to biologic therapies in inflammatory bowel disease. Cell host and microbe, 29(8), 1294–1304. 10.1016/j.chom.2021.06.019

14. Liu, Z., Geboes, K., Colpaert, S., D’Haens, G. R., Rutgeerts, P., and Ceuppens, J. L. (2000). IL-15 is highly expressed in inflammatory bowel disease and regulates local T cell-dependent cytokine production. The Journal of Immunology, 164(7), 3608–3615. 10.4049/jimmunol.164.7.3608

15. Li, X., Conklin, L., and Alex, P. (2008). New serological biomarkers of inflammatory bowel disease. World journal of gastroenterology: WJG, 14(33), 5115. 10.3748/wjg.14.5115

16. McKinney, W. (2010). Data structures for statistical computing in python. In Proceedings of the 9th Python in Science Conference (Vol. 445, No. 1, pp. 51–56).

17. Mehta, R. S., Mayers, J. R., Zhang, Y., Bhosle, A., Glasser, N. R., Nguyen, L. H., et al. (2023). Gut microbial metabolism of 5-ASA diminishes its clinical efficacy in inflammatory bowel disease. Nature medicine, 29(3), 700–709. 10.1038/s41591-023-02217-7

18. Mishra, N., Aden, K., Blase, J. I., Baran, N., Bordoni, D., Tran, F., et al. (2022). Longitudinal multi-omics analysis identifies early blood-based predictors of anti-TNF therapy response in inflammatory bowel disease. Genome medicine, 14(1), 110. 10.1186/s13073-022-01112-z

19. Noor, N. M., Sousa, P., Paul, S., and Roblin, X. (2022). Early diagnosis, early stratification, and early intervention to deliver precision medicine in IBD. Inflammatory bowel diseases, 28(8), 1254–1264. 10.1093/ibd/izab228

20. Piotrowska, M., Spodzieja, M., Kuncewicz, K., Rodziewicz-Motowidło, S., and Orlikowska, M. (2021). CD160 protein as a new therapeutic target in a battle against autoimmune, infectious and lifestyle diseases. Analysis of the structure, interactions and functions. European journal of medicinal chemistry, 224, 113694. 10.1016/j.ejmech.2021.113694

21. Preto, A. J., Chanana, S., Ence, D., Healy, M. D., Domingo-Fernández, D., and West, K. A. (2025). Multi-omics data integration identifies novel biomarkers and patient subgroups in inflammatory bowel disease. Journal of Crohn’s and Colitis, 19(1), jjae197. 10.1093/ecco-jcc/jjae197

22. Schirmer, M., Stražar, M., Avila-Pacheco, J., Rojas-Tapias, D. F., Brown, E. M., Temple, E., et al. (2024). Linking microbial genes to plasma and stool metabolites uncovers host-microbial interactions underlying ulcerative colitis disease course. Cell host & microbe, 32(2), 209–226. 10.1016/j.chom.2023.12.013

23. Seabold, S., and Perktold, J. (2010). statsmodels: Econometric and statistical modeling with Python. In 9th Python in Science Conference.

24. Selin, K. A., Hedin, C. R., and Villablanca, E. J. (2021). Immunological networks defining the heterogeneity of inflammatory bowel diseases. Journal of Crohn’s and Colitis, 15(11), 1959–1973. 10.1093/ecco-jcc/jjab085

25. Souza, H. S., Tortori, C. J., Castelo-Branco, M. T., Carvalho, A. T. P., Margallo, V. S., Delgado, C. F., et al. (2005). Apoptosis in the intestinal mucosa of patients with inflammatory bowel disease: evidence of altered expression of FasL and perforin cytotoxic pathways. International journal of colorectal disease, 20(3), 277–286. 10.1007/s00384-004-0639-8

26. Raffals, L. E., Saha, S., Bewtra, M., Norris, C., Dobes, A., Heller, C., et al. (2022). The development and initial findings of a study of a prospective adult research cohort with inflammatory bowel disease (SPARC IBD). Inflammatory bowel diseases, 28(2), 192–199. 10.1093/ibd/izab071

27. Raine, T., Bonovas, S., Burisch, J., Kucharzik, T., Adamina, M., Annese, V., et al. (2022). ECCO guidelines on therapeutics in ulcerative colitis: medical treatment. Journal of Crohn’s and Colitis, 16(1), 2–17. 10.1093/ecco-jcc/jjab178

28. Virtanen, P., Gommers, R., Oliphant, T. E., Haberland, M., Reddy, T., Cournapeau, D., et al. (2020). SciPy 1.0: fundamental algorithms for scientific computing in Python. Nature methods, 17(3), 261–272. 10.1038/s41592-019-0686-2

29. Waskom, M. L. (2021). Seaborn: statistical data visualization. Journal of open source software, 6(60), 3021. 10.21105/joss.03021

30. Wojtal, K. A., Eloranta, J. J., Hruz, P., Gutmann, H., Drewe, J., Staumann, A., et al. (2009). Changes in mRNA expression levels of solute carrier transporters in inflammatory bowel disease patients. Drug Metabolism and Disposition, 37(9), 1871–1877. 10.1124/dmd.109.027367

