## Supplementary File for "A biomarker catalog for inflammatory bowel disease medications"

#### Supplementary Text

**Supplementary Text 1. Medication Groups:**

In our dataset, we have a total of 18 unique drugs **(Table 1 in the main manuscript)**. When considering these drugs individually, we often end up with sample sizes that hinder the downstream statistical analysis. As an alternative, we grouped the drugs into medication groups, thus improving the statistical significance of the study while maintaining biological plausibility (Table 1). As previously mentioned, the reasoning behind the groupings of these drugs into "medication groups" was centered around their mechanism of action (MOA). We leveraged information on drugs that share an MOA or target(s), such as TNF-α inhibitors.

---

**Supplementary Text 2. Transcriptomic Data Preprocessing:**

Starting from the preprocessed data provided by the SPARC IBD cohort **(see Supplementary Text 1 in Preto et al., 2025)**, we dropped transcripts without a gene name. Using PyDESeq2 (Muzellec *et al*., 2023), we normalized each of the batches. We divided by the size factors (i.e., related to the number of reads in the library), thus making the expression values independent of the number of reads. We also removed features with zero variance (i.e., the same value across all samples) and those with less than 20 total counts across all samples. For transcripts with the same gene name, we averaged the values of the multiple columns. From 38,359 initial features, after this process, the number of unique transcriptomics features is 19,413.

After visualizing the data with the first two components of a Principal Component Analysis (PCA), we identified a steep effect between batches. We performed batch correction using PyComBat-seq (via the inmoose package), and upon visualizing after this, the batch effect was no longer the main separation driver (Behdenna *et al*., 2023). The samples were clustered by tissue, which indicates that these should be modelled separately.

**References:**


- Preto, A. J., Chanana, S., Ence, D., Healy, M. D., Domingo-Fernández, D., and West, K. A. (2025). Multi-omics data integration identifies novel biomarkers and patient subgroups in inflammatory bowel disease. *Journal of Crohn's and Colitis*, *19*(1), jjae197. https://doi.org/10.1093/ecco-jcc/jjae197

- Behdenna, A., Colange, M., Haziza, J., Gema, A., Appé, G., *et al.* (2023). pyComBat, a Python tool for batch effects correction in high-throughput molecular data using empirical Bayes methods. *BMC bioinformatics*, *24*(1), 459. https://doi.org/10.1186/s12859-023-05578-5

- Muzellec, B., Teleńczuk, M., Cabeli, V., and Andreux, M. (2023). PyDESeq2: a Python package for bulk RNA-seq differential expression analysis. *Bioinformatics*, 39(9), btad547. https://doi.org/10.1093/bioinformatics/btad547


---

**Supplementary Text 3. Pathway analysis:** To assess how the gene sets we identify fit into a broader picture, we perform pathway analysis by running them through an Overrepresentation Analysis (ORA) hypergeometric test. We define the gene background as the unique number of transcripts or protein features (respectively) and run them through a Gene Ontology (GO) database.

---

### Supplementary Figures

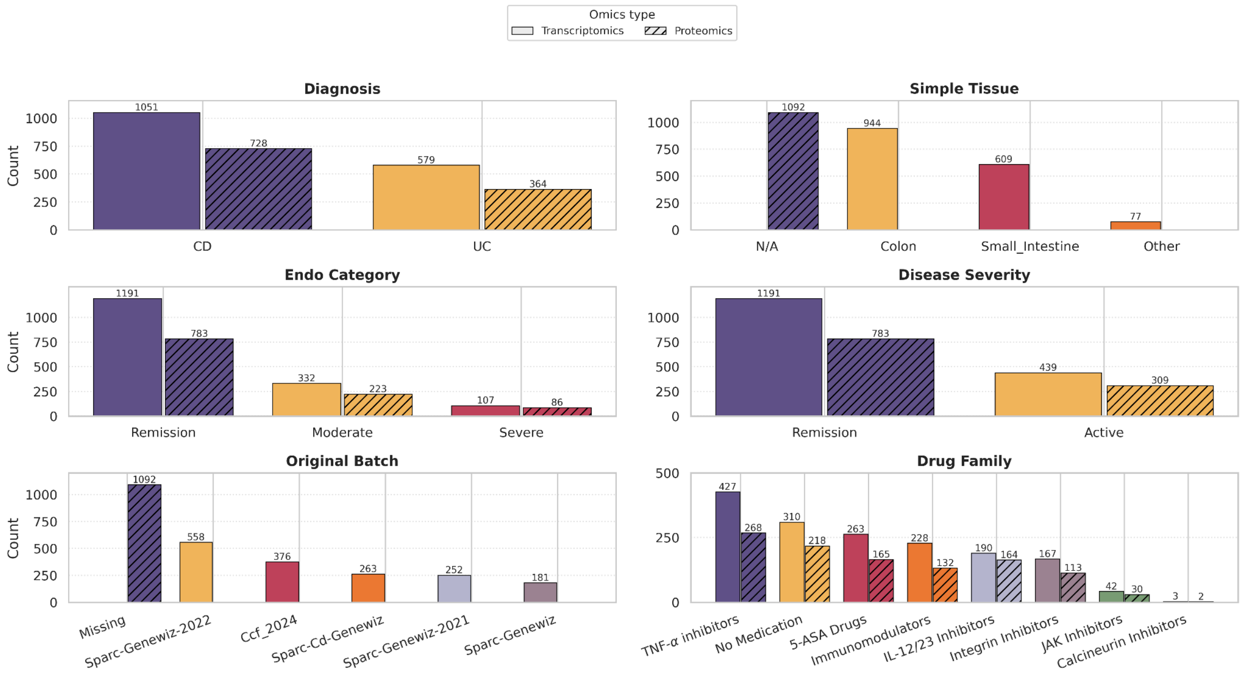


**Supplementary Figure 1. Distribution of relevant metadata information in the samples used for this study.**

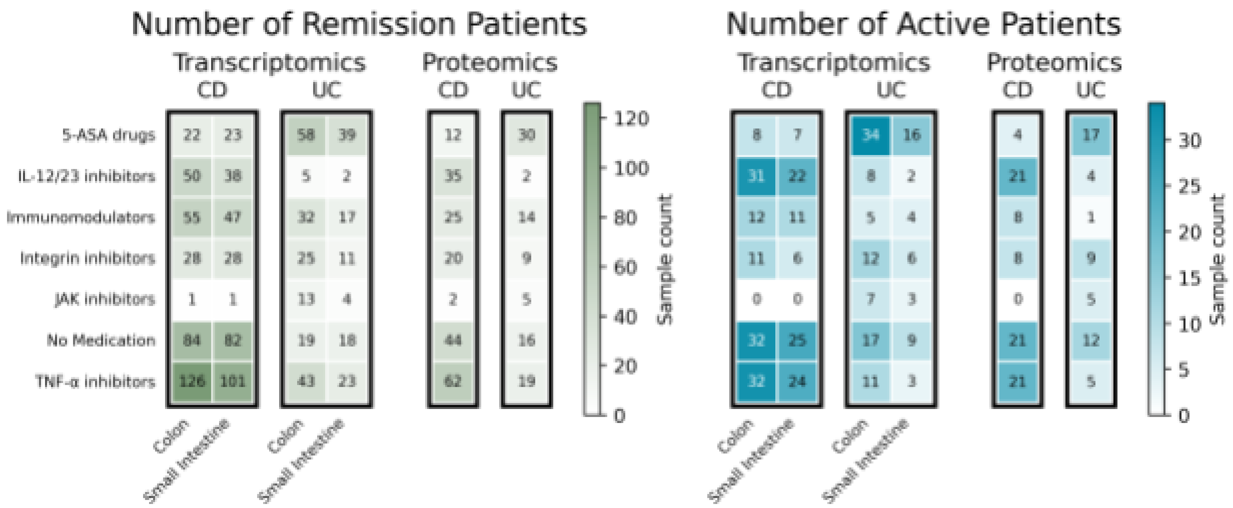


**Supplementary Figure 2. Distribution of patients per subpopulation and medication type split by remission and active disease severity.**

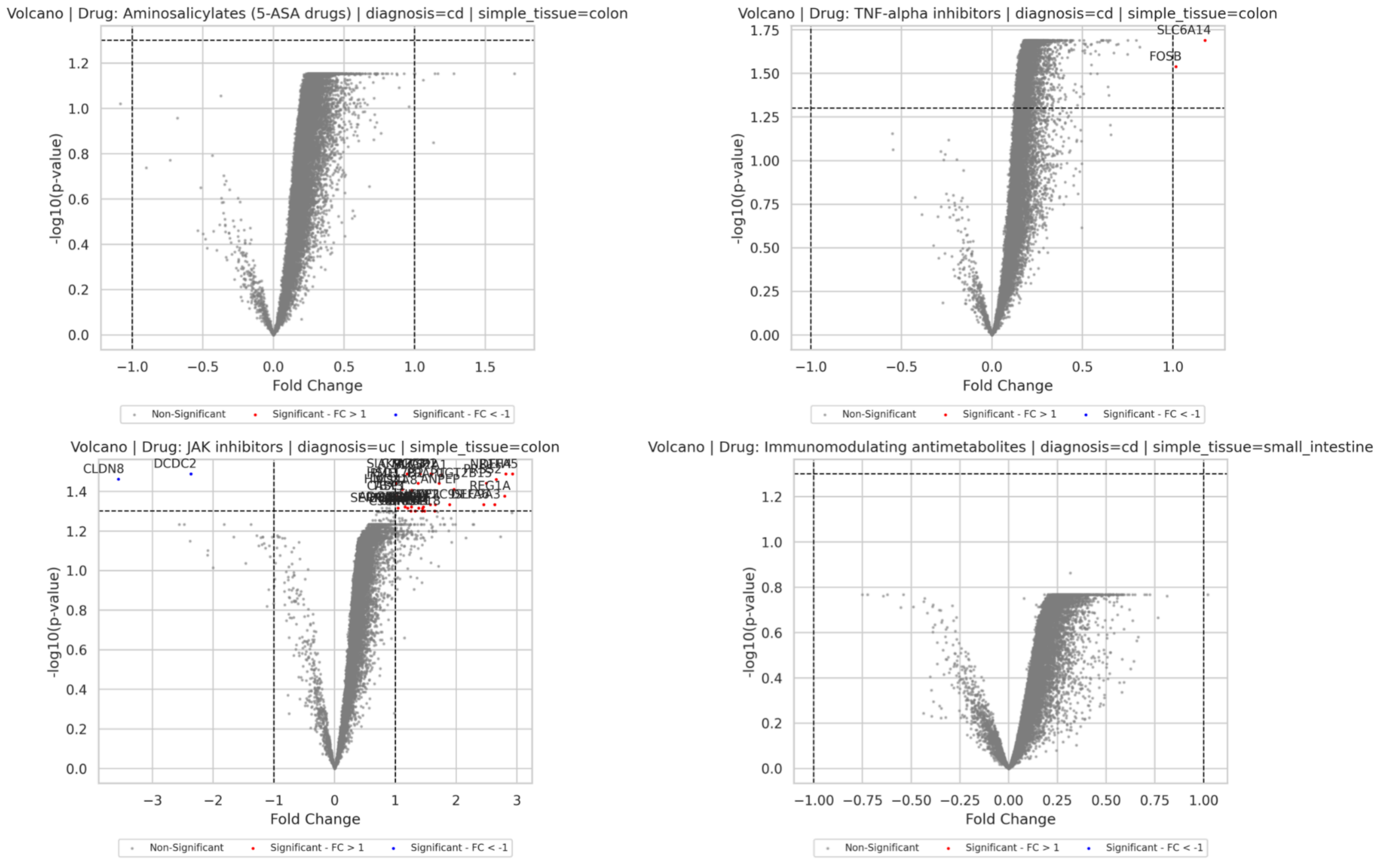


**Supplementary Figure 3. Volcano plots showing more up-regulated transcripts than down-regulated, in various subpopulations for different medication types.**

### Supplementary Tables

| Variable | Explanation |
|---|---|
| Diagnosis | The patient's diagnosis for the provided sample can be categorized as Crohn's Disease (CD), Ulcerative Colitis (UC), or "other/none." We did not consider the "other/none" category in our analysis. |
| Simple Tissue | This refers to the origin of the omics data. Proteomics samples, however, were exclusively taken from plasma and are therefore not associated with specific tissues. For other omics data, biomaterial was categorized into two primary regions: "colon" and "small_intestine." Any regions outside these two, which constituted a minority of samples, were disregarded. |
| Disease Severity | The endo category is derived from a specialized rating system that assesses a patient's disease based on various factors, including Mayo Scores. Disease severity was determined from the Endo Category by combining moderate and mild patients, as detailed in section 2.4 of the main paper. |
| Original Batch | These batches are associated with the cohorts from which the transcriptomic data was obtained. |
| Drug Family | Medications were categorized based on their Mechanism of Action (MOA) or target groups, as detailed in section 2.2 and supplementary text 1. |

**Supplementary Table 1. Metadata employed in the study.**

| | Remission | Mild | Moderate | Severe |
|---|---|---|---|---|
| **Starting** | Tx = 1,847<br>Px = 1,174 | Tx = 1,151<br>Px = 699 | Tx = 635<br>Px = 398 | Tx = 260<br>Px = 173 |
| **Step 1: Filtering out NaN Medications** | Tx = 1,694<br>Px = 1,143 | Tx = 1,046<br>Px = 683 | Tx = 577<br>Px = 384 | Tx = 244<br>Px = 171 |
| **Step 2:<br>After Filtering out “Combination Therapy” and NaN Medication Groups** | Tx = 1,248<br>Px = 813 | Tx = 711<br>Px = 475 | Tx = 369<br>Px = 253 | Tx = 131<br>Px = 92 |
| **Step 3 :** | Tx = 1191 | Tx = 0 | Tx = 332 | Tx = 107 |

| | | | | |
|---|---|---|---|---|
| **Filtering down to just remission and active (moderate, severe) patients and removing patients with samples on different Medications** | Px = 783 | Px = 0 | Px = 223 | Px = 86 |

**Supplementary Table 2. Sample counts at each step of data cleaning.**

### Figure Credits:

Magazine free icon by Nadiinko: Flaticon.com
